# Brain functional connectivity and growth measurements in near-term and term-born neonates: an fNIRS study

**DOI:** 10.64898/2026.05.14.26349878

**Authors:** Cassia Donga, Lingkai Tang, Kerlas Samaan, Kevin Stubbs, Homa Vahidi, Soume Bhattacharya, Christine Grafe, Sandrine de Ribaupierre, Keith St. Lawrence, Emma G. Duerden

## Abstract

Resting-state networks (RSNs), measured through functional connectivity (FC), emerge in utero and are detectable within hours of birth. Although neonatal growth metrics predict later neurodevelopmental outcomes and structural brain maturation, their relationship to early functional network organization remains poorly understood. We examined associations between anthropometric growth metrics and resting-state FC in a cohort of healthy near-term and term-born neonates using functional near-infrared spectroscopy (fNIRS) acquired during the first few days of life. Task-free fNIRS data were recorded in 121 neonates (67 males [55%]; mean postnatal age=25.6 hours; mean gestational age=38.63 weeks). Based on birthweight percentiles, 12 (9%) newborns were small for gestational age (SGA) and 13 (11%) were large for gestational age (LGA). Growth metrics included birth weight–for–gestational age z-score (BGZ), head circumference–for–gestational age z-score (HGZ), birth weight–for–length z-score (BLZ), and z-scored Ponderal Index (PI-z). Whole-brain FC was calculated as the mean Fisher-Z-transformed correlation across valid channel pairs. Channel-wise associations were examined using general linear and linear mixed-effects models controlling for gestational age, postnatal age and sex. Linear and quadratic terms were tested, and multiple comparisons were controlled using the false discovery rate. None of the anthropometric measures were associated with global FC; however, significant nonlinear (quadratic) relationships emerged at the channel-pair level. BGZ (β range= −0.102 to −0.074; FDR-corrected p<0.005) and PI-z (β range= −0.088 to −0.074; FDR-corrected p < 0.001) demonstrated negative quadratic associations with inter- and intrahemispheric connectivity, such that newborns with both lower (SGS) and higher (LGA) growth values showed reduced FC relative to those with average growth. In contrast, HGZ demonstrated positive quadratic associations (β range=0.051 to 0.074; FDR-corrected p<0.001), with infants at the lower and higher ends of the head-size distribution exhibiting increased FC relative to infants near the mean. BLZ showed no significant associations after correction. Results indicate that early somatic growth is reflected in the organization of neonatal functional brain networks and that deviations from average growth whether smaller or larger are associated with altered regional connectivity. Findings suggest that neonatal growth metrics may provide an accessible marker of early brain health, reflected in regionally specific functional connectivity patterns.

## 1.0 Introduction

Early brain development is marked by the establishment of various cognitive functions, of which an important indicator is the early maturation of functional connectivity (FC). In newborns, functional networks emerge *in utero* and continue to evolve rapidly during the first weeks of life.^1,2^ These networks, typically observed through functional magnetic resonance imaging (fMRI), have been more recently examined using functional near-infrared spectroscopy (fNIRS).^3–5^ Due to its portability, low cost and high tolerance for motion, compared to fMRI, fNIRS has become a valuable tool for studying neonatal brain FC, a key marker for brain health.^6,7^

Existing fNIRS research demonstrates that newborns exhibit developing networks in frontal, temporal, and parietal regions.^5^ Altered fNIRS-based connectivity has also been observed in preterm infants with perinatal complications^8,9^ indicating that early functional networks are sensitive to clinical and developmental factors. fNIRS-based FC measures relate to behavioural traits such as temperament and regulatory functioning,^8,10^ supporting the interpretation that early functional organization carries cognitive significance.

Importantly, neurovascular coupling is still developing in preterm and term newborns, resulting in immature and sometimes inverted hemodynamic responses. Unlike adults, newborns commonly exhibit delayed, attenuated or even negative hemodynamic response patterns, with substantial regional variability. These developmental differences highlight the need to interpret neonatal FC within a physiological framework that accounts for heterogeneous vascular maturation.^11^

In parallel with neural maturation, newborn body growth reflects the adequacy of prenatal and early postnatal nutrition, metabolic stability, and environmental conditions. Deviations from expected growth trajectories, such as small for gestational age (SGA) or large for gestational age (LGA), are associated with later differences in cognitive ability, academic skills and neurodevelopmental outcomes.^12–17^ These relationships underscore the importance of physical growth as an early marker of developmental risk.

Although body growth and brain FC, as two vital indicators of the health of neonates, are often studied separately, increasing evidence suggests that they are strongly interrelated. For instance, preterm-birth induced low body weight is commonly associated with abnormal functional or structural connectivity.^18^ However, relatively few studies have focused on FC and anthropometric measurements (AMs) in the term-born population, to establish normative trajectories.

This study aimed to investigate the relationship between FC, measured using fNIRS and AMs in term-born neonates. In a single centre cohort study of newborns scanned with fNIRS during the first few days of life, we investigated linear and nonlinear relationships between multiple anthropometric indicators including birth weight–for–gestational age z-score (BGZ), head circumference–for–gestational age z-score (HGZ), birth weight–for–length z-score (BLZ), and the Ponderal Index (PI-z) for both global and regional connectivity patterns.

We hypothesized that neonatal body growth would be associated with fNIRS-based resting-state FC. Specifically, we expected that infants with lower or higher growth values such as those SGA or LGA would show altered FC relative to infants with average growth. Based on prior evidence that neonatal neurovascular and physical development do not follow strictly linear patterns,^19^ we further hypothesized that these associations would include nonlinear (quadratic) effects and would show regionally-specific associations rather than global changes across the cortex. By characterizing how early functional network organization relates to measures of neonatal growth, our goal was to identify potential markers of typical versus atypical developmental trajectories in the first days of life.

## 2.0 Methods

### 2.1 Study Design & Participants

This was a prospective cohort study of healthy near-term and term born neonates within the first hours of birth. Participants were recruited between 2020-2025. This study was approved by the Western University Health Sciences Research Ethics Board (REB #116142), and verbal and written consent was obtained from all legal guardians.

Participants included neonates being cared for in the Post Partum Care Unit (PPCU) at a large obstetrical and pediatric hospital in Southwestern Ontario. Neonates were eligible to participate if they had a gestational age of at least 35+0 weeks and were deemed healthy by the attending pediatrician. Neonates were not considered eligible if they met any of the following criteria: congenital malformation or syndrome, antenatal exposure to illicit drugs, postnatal infection, and suspected brain abnormalities and/or injuries. A member of the patient’s circle of care was first involved to determine eligibility and the family’s potential interest in participating. The circle of care members approached the infants’ legal guardians to inquire if they were comfortable being approached by a member of the research team to learn more about participation. A member of the research team then approached interested families and provided all relevant information, time to ask questions and to consider their decision as needed. All caregivers provided written informed consent. The study procedures were carried out according to the Declaration of Helsinki.

Once informed consent was obtained, the infants head circumferences were obtained from the newborns’ charts and were subsequently used to populate the appropriately sized mesh cap. Demographic data was obtained from charts by nurses including the infant’s exact gestational age, time of birth, birth weight, head circumference, crown–to–heal length, cord clamping, Apgar’s scores, anesthesia, mode of delivery and resuscitation at delivery. Nurses also collected demographic data from the mother’s charts including maternal age, serologies, comorbidities, blood type and antibodies. The most recent feeding time was verbally confirmed with the infant’s legal guardian prior to recordings.

### 2.2 Data Collection

To obtain fNIRS recordings, mesh cap (Easycap GmbH, Germany) sizes were matched to the head circumference measured at birth and recorded in the chart. The Easycap’s used are made in even sizes, so for infants with odd-numbered HCs, we chose to round up to the nearest even number, for comfort. Caps were then populated and placed on the infants’ heads, assuring proper positioning by measuring nasion-to-inion distance to center Cz on the anterior-posterior axis and measuring from the left to right ear canal to center Cz along the sagittal plane. Data was most frequently collected from infants after feeding and when calm to decrease fussiness and motion, when possible.

Resting-state, task-free fNIRS data was collected in the neonates at bedside using a multichannel NIRSport2 system (Aurora Software, NIRx Medical Technologies LLC, Berlin, Germany) with a sampling rate of 10.17 Hz. The montage included 10 channels per hemisphere, made up of 8 LED sources (760 and 850 nm) and 8 detectors, with an average source-detector separation of 35.7 ± 9.5 mm (Figure 1). Previous literature recommended sampling time to include a minimum of 2.5 minutes of recording.^20^ However, to ensure enough representative data was collected, we recorded for a minimum of 10 minutes, when possible, with recordings lasting up to 16 minutes if the infant was comfortable. Recordings were all started when the infants were awake and alert with their eyes open, however without confirmation via electroencephalography, we cannot be certain if each infant remained awake for the entire duration of the recordings. Behavioral state (e.g., quiet sleep vs wakefulness) was not formally coded and therefore not included as a covariate in any of the subsequent analyses. Data collection was stopped if the infant was moving too much to keep the cap in place, inconsolable or at any time if requested by the legal guardian. All demographic and fNIRS data were stored on a password locked computer, in a restricted-access area at on the care unit.

**Fig 1.**
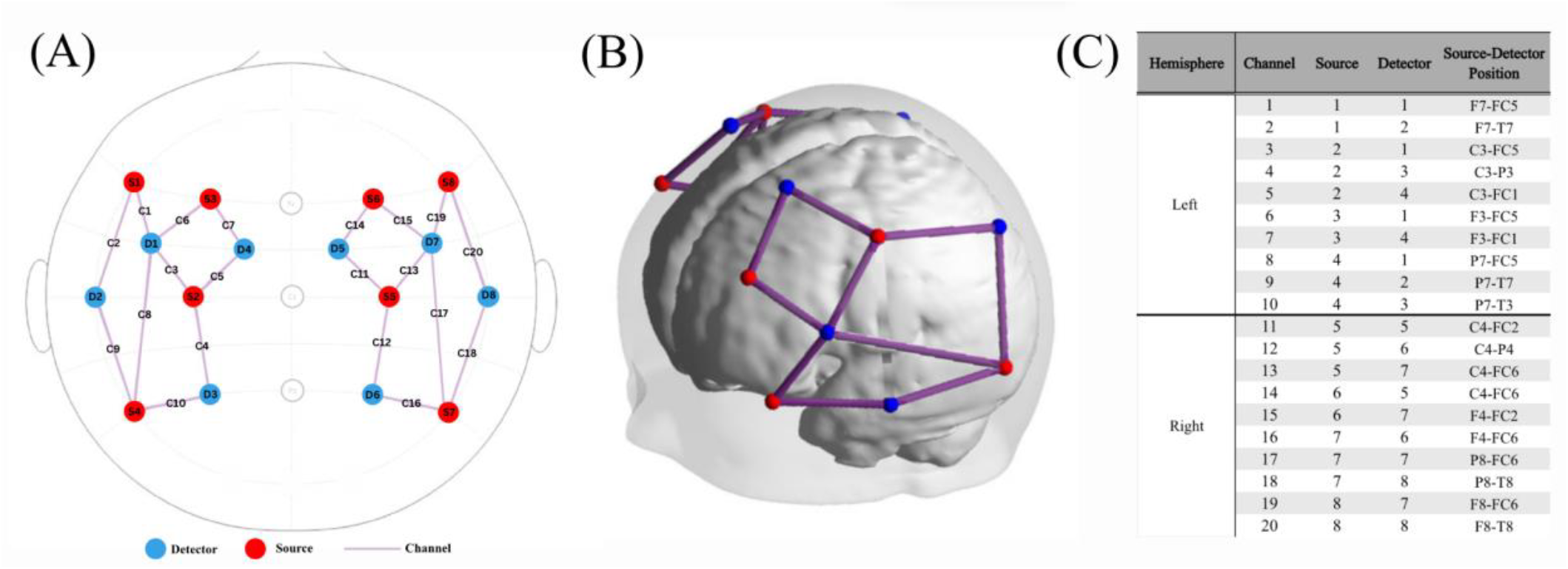
(A) Infant montage including 8 sources and 8 detectors placed on a 10-10 system cap. (B) 3-Dimensional view of source-detector placement. (C) Exact placement of channels made up from one source and one detector each, on a 10-10 system.

Three infants were excluded from preprocessing due to lost data, resulting in 121 neonates with complete recordings. This cohort comprised 67 males, 54 females and 12 infants that were deemed SGA by their pediatrician, with a mean gestational age of 38.63 ± 1.39 weeks and a mean postnatal age of 25.6 ± 13.89 hours. Of the 121 infants, 57 were born by vaginal birth and 64 were born via Cesarean section.

**Table 1.** Demographic data.

|  | Mean | SD |
| --- | --- | --- |
| Postnatal age | 25.6 hours | 13.89 |
| Gestational age | 38.63 weeks | 1.39 |
| Birthweight | 3211.21 g | 618.46 |
| Head circumference | 34.67 cm | 2.5 |
| Length | 49.69 cm | 3.62 |

Cm, centimeters, g, grams, SD, standard deviation

### 2.3 Statistical Analysis

#### 2.3.1 Normality of Anthropometric Measures

Analyses were conducted using SPSS (Version 29). All anthropometric variables were analyzed using gestational age–adjusted z-scores, consistent with the World Health Organization (WHO) Multicentre Growth Reference Study Group (2006), the WHO (2004)^21^ and recommended neonatal growth modelling practices. As these standardized scores are designed to approximate a normal distribution, we evaluated normality using the Shapiro–Wilk test. Outliers were defined as values exceeding ±3 SD and were removed prior to statistical analysis.

#### 2.3.2 Growth Parameters

Various growth parameters, including BGZ, HGZ, BLZ, and the Ponderal Index, were calculated for each infant using Microsoft Excel (Microsoft Corporation, 2024) based on recorded AMs. BGZ was derived by comparing each infant’s birth weight to the sex-specific expected birth weight– for–gestational age using the World Health Organization Child Growth Standards. HGZ was calculated by comparing each infant’s head circumference to the corresponding gestational age- and sex-specific reference values from the World Health Organization Growth Standards BLZ was calculated using the LMS method, incorporating gestational age-specific reference values for birth length as defined by the World Health Organization Growth Standards. The Ponderal Index was calculated as birth weight divided by birth length cubed (kg/m^3^) to provide an index of neonatal body proportionality. Additionally, the Ponderal Index was z-scored (PI-z) to be used in modelling. Each growth parameter was assessed for outliers, with those greater than 3 SDs from the mean, being removed. Further, this resulted in the removal of 3 participants for HGZ, 4 participants for BLZ and one participant for PI-z.

Classifications of babies considered SGA, average for gestational age (AGA) or LGA were determined separately for each measurement. Using the z-score of each growth parameter, babies with a z-score smaller than -1.28 (the 10th percentile on a z-scale) were categorized as SGA for that growth parameter. Infants with a z-score larger than 1.28 (the 90th percentile on a z-scale) were categorized as LGA. All other infants were categorized as AGA for each respective growth parameter.

#### 2.3.3 Preprocessing Pipeline

The fNIRS data underwent preprocessing through a multi-step pipeline executed in MATLAB (MathWorks) utilizing the AnalyzIR toolbox. This approach was adapted from an established framework for motion correction and resting-state processing,^22^ as illustrated in Figure 2. All analyses were conducted utilizing continuous-wave intensity data collected at wavelengths of 760 and 850 nm.

**Figure 2.**
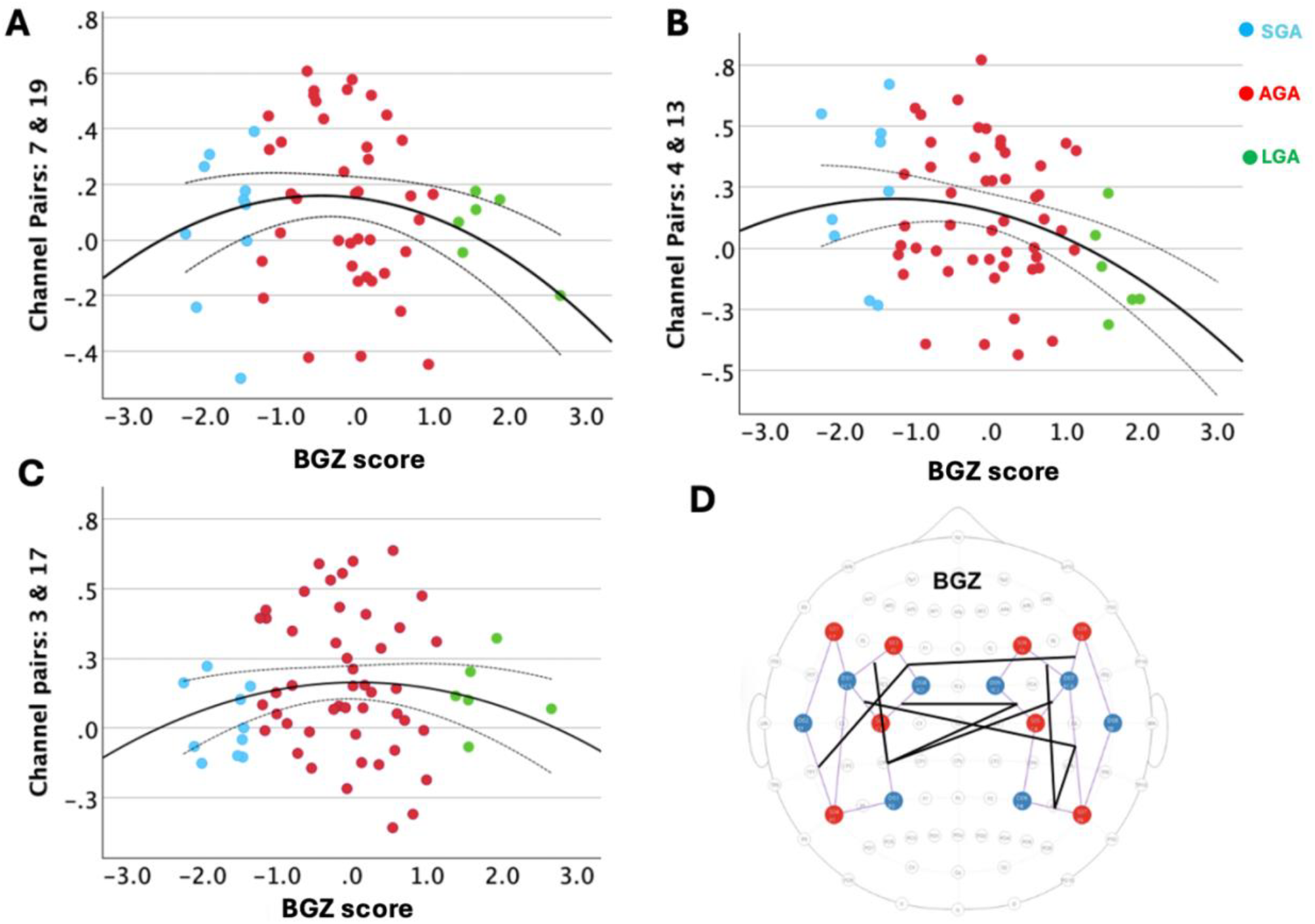
Quadratic associations between BGZ and resting-state FC in neonates. (A–C) Scatterplots showing significant quadratic relationships between BGZ and FC for three channel pairs (A: 7–19; B: 4–13; C: 3–17). Higher or lower BGZ values were associated with reduced FC relative to infants with average BGZ. Solid lines indicate the fitted quadratic function; dotted lines show the 95% confidence interval. Individual data points represent HbO-based FC values for each infant. (D) Spatial representation of significant channel-pair associations displayed on the 20-channel fNIRS sensor layout. Red and blue nodes denote channels contributing to significant effects; black connecting lines identify inter- and intra-hemispheric channel pairs surviving FDR correction.

##### 2.3.3.1 Assessment of quality metrics and the selection process for clean data segments

Initially, we implemented Temporal Derivative Distribution Repair (TDDR) on the raw intensity signals to address baseline shifts and mitigate the majority of motion spikes.^22^ This was subsequently followed by wavelet filtering utilizing a fixed interquartile range (IQR) threshold of 1.5 to reduce any remaining motion artifacts.^23^ For each infant, the cardiac frequency range was determined utilizing the Fourier spectrum derived from motion-corrected signals or, when required, through the examination of unblemished time segments within the raw traces. In this customized cardiac band, we calculated the scalp coupling index (SCI) and peak spectral power (PSP) utilizing a 3-second sliding window. Samples were categorized as clean when the SCI value exceeded 0.7 and the PSP value surpassed 0.1.^24,25^ Participants with ≥30% of clean channels, based on the previously calculated SCI and PSP, were excluded entirely from the subsequent step window calculation step. This threshold was selected to balance data retention with signal reliability, ensuring sufficient spatial coverage for connectivity estimation while accommodating the high motion susceptibility and variable scalp coupling characteristic of neonatal fNIRS recordings.^26^ Nine infants failed to satisfy this ≥30% clean-channel inclusion criterion, resulting in a final sample size of 112 infants (50 females, 62 males) for the FC analyses.

For each dataset, we subsequently identified the 300-second window from the raw data that contained the highest quantity of clean samples. In instances where multiple candidate windows exhibited equal counts, the earliest window was chosen for analysis. Channels were omitted from subsequent analysis if they exhibited less than 70% clean samples within the designated 300-second segment (SCI ≤ 0.7 and PSP ≤ 0.1). Excluded channels were retained as placeholders, populated with constant values, to maintain the integrity of matrix dimensions, and were subsequently assigned NaN during the correlation phase. Infants for whom 70% or more of channels were classified as poor quality within the selected segment were excluded from subsequent connectivity analyses, consistent with the participant-level quality control criterion described above, resulting in the exclusion of nine infants (5 males and 4 females).

##### 2.3.3.2 Optical density and hemodynamic conversion

Following the selection of the 300-second resting-state segment and the exclusion of low-quality channels, the intensity data underwent conversion to optical density (OD). The TDDR methodology was implemented a second time on optical density signals, subsequently followed by the application of wavelet filtering techniques. In the second wavelet step, we conducted a systematic evaluation of interquartile range thresholds ranging from 1.0 to 1.5, incrementing by 0.1. For each individual infant, we identified the threshold that most effectively eliminated observable motion artifacts throughout the subsequent processing stages. The optical density data underwent a transformation to quantify concentration variations in oxy- and deoxy-hemoglobin (HbO, HbR). This process utilized the modified Beer-Lambert Law. This analysis incorporates differential pathlength factors that are appropriate for the age of the subjects involved and channel distances scaled to the measured head circumference in a 0-to 2-month infant template.

##### 2.3.3.3 Frequency filtering and connectivity metric

Hemoglobin time series underwent bandpass filtering within the frequency range of 0.009 to 0.08 Hz, utilizing MATLAB’s bandpass function, which employs a minimum-order Infinite Impulse Response (IIR) filter characterized by a 60 dB stopband attenuation.^27^ Zero padding was utilized during filtering. This procedure was implemented to isolate low-frequency resting-state fluctuations while effectively minimizing edge artifacts. The total hemoglobin (HbT) was calculated by summing the concentrations of HbO and HbR, serving as the principal signal for the connectivity analysis.^28,29^ HbT was selected as the primary signal for connectivity analyses because it is less sensitive to polarity inversions and regional variability in neurovascular coupling commonly observed in neonates

The nirs.sFC.ar_corr function was used to estimate resting-state functional connectivity (FC). This function uses a strong Pearson correlation method with pre-whitening to address temporal autocorrelation in the narrow-band signal.^30^ The selection of model order was conducted utilizing the Bayesian Information Criterion, with a maximum limit set at 51, which corresponds to five times the sampling rate.^31^ All subsequent statistical analyses involved the Fisher-Z transformation of correlation coefficients into Z values.

#### 2.3.4 Total Functional Connectivity

Total FC was computed for each infant by averaging FC values across all valid channel pairs. To examine associations between global connectivity and AMs, we ran a series of general linear models (GLMs). Each model included a single anthropometric predictor (BGZ, HGZ, BLZ, or PI-z) to avoid multicollinearity among growth indices. All models controlled for postnatal age at testing, gestational age at birth, and sex. Both linear and quadratic terms were tested to capture potential nonlinear (inverted-U) relationships between FC and AMs.

#### 2.3.5 Channel-wise Comparisons

To assess spatially specific associations between growth parameters and FC, we conducted channel-wise linear mixed-effects (LME) models. Separate models were estimated for each channel pair, with FC as the dependent variable and each AM as a predictor. Sex and postnatal age were included as fixed covariates, and a random intercept for participants was included to account for within-infant dependence across channels. Both linear and quadratic terms were evaluated. P-values were corrected for multiple comparisons across all channel pairs using the false discovery rate (FDR).

## 3. Results

### 3.1 Participants

Verbal and written consent was obtained from the legal guardians of 131 infants, 7 of which, withdrew consent prior to recordings, due to changes in the infant’s health status or general fussiness. Resting-state, task-free fNIRS data was collected in 124 neonates. With exclusions, a total of 121 healthy neonates (67 males [55%], 54 females) born at ≥35+0 weeks gestation were included in the analysis. Delivery mode included 57 (47%) vaginal births and 64 (53%) caesarean deliveries.

Anthropometric measures included: BGZ, HGZ, BLZ, and PI-z. Based on birthweight percentiles, 12 (9%) newborns were SGA and 13 (11%) were LGA.

### 3.2 Whole-Brain Functional Connectivity

Generalized linear models revealed no significant relationships between any anthropometric measures (BGZ, HGZ, BLZ, or PI-z) and whole brain mean FC derived from HbT correlations (all, p>0.05).

### 3.3 Channel-wise Functional Connectivity

#### 3.3.1 Nonlinear Associations with BGZ

Linear mixed effects models revealed significant negative (U-shaped) quadratic associations between BGZ and channel-wise FC across multiple cortical connections (β = −0.102 to −0.074, FDR-corrected p<0.005). As shown in infants with both lower and higher BGZ (i.e., SGA and LGA) demonstrated *reduced* FC compared to those with average BGZ, consistent with an inverted-U pattern (Figure 2A–C). These effects were not global but instead demonstrated a distinct spatial organization. The channel-mapping representation (Figure 2D) shows that significant BGZ–FC associations were distributed across frontal, temporal, and parietal regions, spanning both inter-hemispheric (e.g., left–right frontal and temporal connections) and intra-hemispheric pathways. No linear associations between BGZ and channel-wise FC survived FDR correction.

#### 3.3.2 Nonlinear Associations with PI

Similarly, PI showed significant negative (U-shaped) quadratic associations with FC (β = −0.088 to −0.074; FDR-corrected p<0.001). These effects mirrored the BGZ pattern, where deviations from average neonatal proportionality corresponded to lower network connectivity. No linear associations between PI and channel-wise FC survived FDR correction.

#### 3.3.3 Nonlinear Associations with Head-Circumference-for-GA (HGZ)

HGZ demonstrated positive (inverted U shaped) quadratic effects on FC across five channel pairs (β = 0.051 to 0.074; FDR-corrected p < 0.001, Figure 3A-B). In turn, newborns at both the low and high ends of the head-size distribution showed increased FC, whereas those near the mean HGZ showed slightly reduced FC. No linear associations between HGZ and channel-wise FC survived FDR correction.

**Figure 3.**
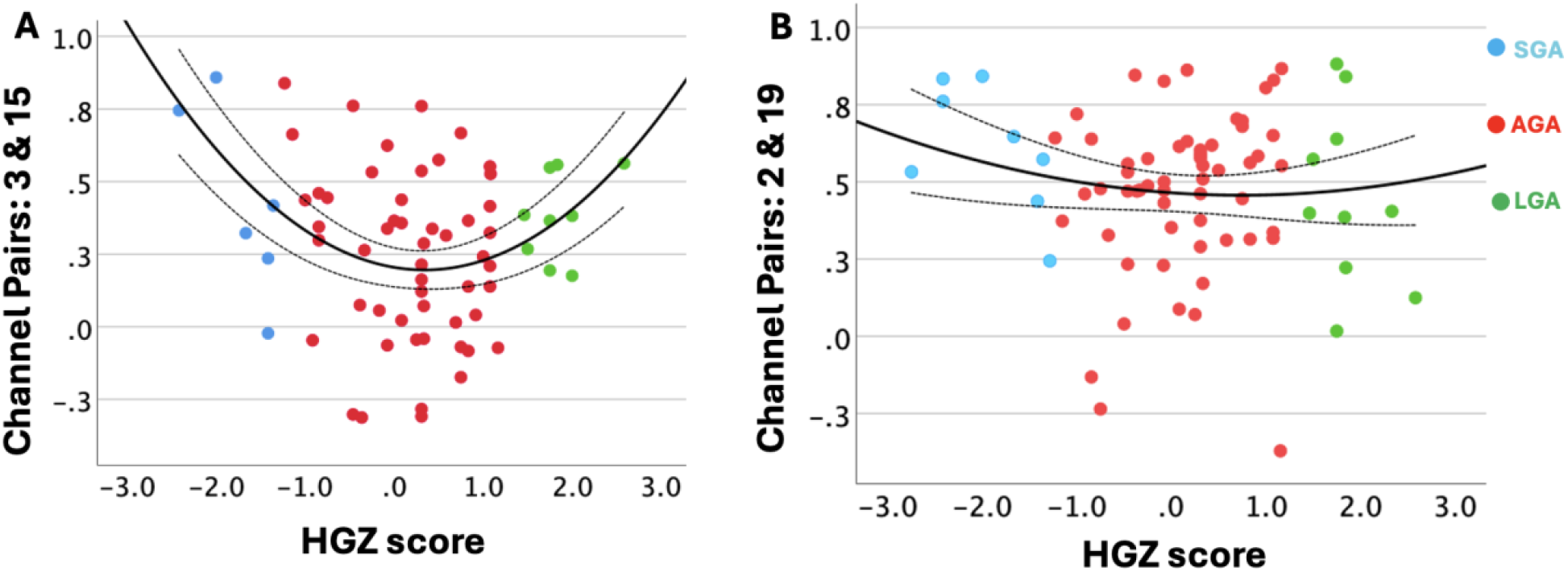
U-shaped associations between head circumference and functional connectivity measured with fNIRS. (A) FC between channel pairs 3–15 and (B) channel pairs 2–19 p lotted as a function of HGZ. Scatter points represent individual participants, with colors indicating group membership. Solid black lines depict the fitted quadratic (U-shaped) regression, with dashed lines indicating 95% confidence intervals. Across both channel pairs, functional connectivity was lowest at average head circumference and increased at both lower and higher HGZ.

As part of an exploratory analysis, we further examined mode of delivery in relation to HGZ and FC, given known influences of vaginal delivery on head circumference, particularly in babies born LGA. Results showed that mode of delivery had borderline effect in these models (p = 0.058) but did not alter the significance of HGZ and FC in the channel pairs.

## 4. Discussion

In a homogeneous cohort of healthy near-term and term-born neonates, we demonstrated that neonatal growth metrics predict variations in resting-state FC within the first days of life. Findings support an association between early physical development and large-scale brain network organization. While prior work has shown that AMs relate to structural brain development, few studies to date have examined their association with functional networks at birth. Findings extend this literature by showing non-linear relationships between key growth indices at birth and connectivity patterns across multiple resting state networks obtained within the first few days of life. Results suggest that even subtle differences in intrauterine growth may shape the functional architecture of the newborn brain. Early growth trajectories should be considered when interpreting neonatal FC measures.

Early growth metrics, particularly head circumference, are well-established indicators of later neurodevelopmental outcomes.^32–35^ Studies with preterm born and growth-restricted neonates consistently show that variations in birth weight and head circumference predict later cognitive performance and brain development.^36^ Slower postnatal growth predicted delayed cortical microstructural development, independent of brain injury or illness in very preterm born neonates.^36^ Further, inadequate post-discharge head growth was associated with lower IQ in very preterm children^37^ and early head growth trajectories strongly predict neurodevelopment into adulthood.^38^ Early brain growth indices, such as corpus callosum length or rapid head circumference gain, provide meaningful predictive value for later functioning.^39,40^ Our findings extend this literature by showing that growth metrics also predict functional network organization at birth, suggesting that early growth contributes to shaping the newborn connectome.

Head circumference and birth weight are commonly used indices of neonatal growth capture partially distinct biological processes.^41,42^ Head circumference primarily reflects prenatal brain growth and cranial development,^41^ whereas birth weight reflects more global somatic growth influenced by placental efficiency, metabolic status, and fluid balance.^43^ The U-shaped association between HGZ and FC may reflect a balance between growth constraint and maturational acceleration. At the lower end of the head circumference distribution, increased connectivity may reflect compensatory or less differentiated network organization in response to constrained brain growth.^44^ At the higher end, increased connectivity may reflect accelerated or advanced network integration associated with larger brain volume and greater synaptic density.^45^ Mode of delivery was only borderline associated with HGZ and is therefore unlikely to fully account for the observed nonlinear associations, although transient perinatal factors related to delivery may contribute modestly to early hemodynamic variability. In contrast, newborns with average head circumference may exhibit more differentiated, selectively organized networks, resulting in comparatively lower global connectivity. The inverted U-shaped association observed for BGZ suggests that intermediate somatic growth may be most conducive to early functional network organization. Both lower and higher birth weight may reflect suboptimal intrauterine conditions, including placental insufficiency at the lower end and altered metabolic or inflammatory environments at the higher end, which may disrupt early functional integration.^46–48^

The observed variability in neonatal functional connectivity, along with the nonlinear associations with growth parameters, is consistent with the immature neurovascular environment of preterm and term infants. At this stage, autoregulatory mechanisms are still developing, leading to hemodynamic responses that are often delayed, attenuated, or inverted relative to adults, a pattern that has been linked to heightened sensitivity to systemic blood pressure and underdeveloped cerebrovascular control. ^11^ Differences in hemodynamic response function (HRF) morphology between preterm and term infants further contribute to regional heterogeneity in functional responses, reflecting distinct maturation trajectories across cortical networks.^49,50^ The timing and shape of the neonatal HRF also diverge from adult patterns, which has important implications for interpreting connectivity estimates in early life.^1,51^ Taken together, these developmental factors likely influence both the magnitude and spatial distribution of connectivity–growth associations observed in our sample, underscoring the need for age-appropriate neurovascular models when examining infant FC.^52^

While this study has a number of strengths including rare high-density fNIRS recordings in hours old newborns, the results should be considered with several limitations. This was a single centre cohort, the sample included a wide gestational age range, but few newborns were SGA or LGA limiting the generalizability of the results to newborns born outside normative ranges for weight, the fNIRS data were acquired largely with the same cap, however magnetic resonance imaging (MRI) was not performed or another method to precisely locate where in the cortex the optodes were being placed. Channel locations were interpreted using a neonatal head template aligned to the international 10–10 system; individual MRI-based registration was not performed, and in turn cortical correspondence should be considered approximate. An additional consideration is that the data were acquired in a cross-sectional design. At our centre, healthy newborns are discharged 24-72 hours from delivery in turn a longitudinal design would not be possible; however, future research could focus on post-discharge scans to further determine the associations between early growth metrics and functional connectivity.

## 5. Conclusions

Across multiple anthropometric indicators, nonlinear (quadratic) relationships emerged between neonatal growth and regional functional connectivity, while global connectivity remained unaffected. Findings indicate that brain–body coupling in the first days of life follows non-linear, regionally specific developmental patterns, with both lower- and higher–than–average growth associated with altered FC relative to normative ranges.

## Data Availability

All data produced in the present study are available upon reasonable request to the authors and with appropriate ethical approval and data transfer agreements.

## References

1. Smyser CD, Snyder AZ, Shimony JS, Mitra A, Inder TE, Neil JJ. Resting-State Network Complexity and Magnitude Are Reduced in Prematurely Born Infants. Cerebral Cortex. Epub 2014.

2. Doria V, Arichi T, Merchant N, et al. Emergence of resting state networks parallels thalamocortical connections formation in the developing brain: Insights from a resting state fMRI study. J Anat. 2011;218.

3. Vahidi H, Kowalczyk A, Stubbs K, et al. Investigating Task-Free Functional Connectivity Patterns in Newborns Using Functional Near-Infrared Spectroscopy. Brain Behav. 2024;14.

4. Hu A, Tong X, Yang L, et al. Gender differences in the functional language networks at birth: a resting-state fNIRS study. Cerebral Cortex. 2024;34.

5. Homae F, Watanabe H, Otobe T, et al. Development of global cortical networks in early infancy. Journal of Neuroscience. 2010;30.

6. Sakai J. Functional near-infrared spectroscopy reveals brain activity on the move. Proc Natl Acad Sci U S A. 2022;119.

7. Hu Z, Liu G, Dong Q, Niu H. Applications of Resting-State fNIRS in the Developing Brain: A Review From the Connectome Perspective. Front. Neurosci. 2020.

8. Fuchino Y, Naoi N, Shibata M, et al. Effects of Preterm Birth on Intrinsic Fluctuations in Neonatal Cerebral Activity Examined Using Optical Imaging. PLoS One. 2013;8.

9. Tang L, Kebaya LMN, Altamimi T, et al. Altered resting-state functional connectivity in newborns with hypoxic ischemic encephalopathy assessed using high-density functional near-infrared spectroscopy. Sci Rep. 2024;14.

10. Kelsey CM, Farris K, Grossmann T. Variability in Infants’ Functional Brain Network Connectivity Is Associated With Differences in Affect and Behavior. Front Psychiatry. 2021;12.

11. Kozberg MG, Chen BR, DeLeo SE, Bouchard MB, C. Hillman EM. Resolving the Transition From Negative to Positive Blood Oxygen Level-Dependent Responses in the Developing Brain. Proceedings of the National Academy of Sciences. Epub 2013.

12. Leitner Y, Fattal-Valevski A, Geva R, et al. Neurodevelopmental outcome of children with intrauterine growth retardation: A longitudinal, 10-Year prospective study. J Child Neurol. 2007;22.

13. Pryor J, Silva P, Brooke M. Growth, development and behaviour in adolescents born small-for-gestational-age. J Paediatr Child Health. 1995;31.

14. McCormick MC, Workman-Daniels K, Brooks-Gunn J. The behavioral and emotional well-being of school-age children with different birth weights. Pediatrics. 1996;97.

15. Ferguson KK, Sammallahti S, Rosen E, et al. Fetal Growth Trajectories among Small for Gestational Age Babies and Child Neurodevelopment. Epidemiology. 2021;32.

16. Eves R, Mendonça M, Bartmann P, Wolke D. Small for gestational age—cognitive performance from infancy to adulthood: an observational study. BJOG. 2020;127.

17. Yu B, Garcy AM. A longitudinal study of cognitive and educational outcomes of those born small for gestational age. Acta Paediatrica, International Journal of Paediatrics. 2018;107.

18. Brenner RG, Wheelock MD, Neil JJ, Smyser CD. Structural and functional connectivity in premature neonates. Semin Perinatol. 2021;45.

19. López-Guerrero N, Alcauter S. Developmental Trajectories and Differences in Functional Brain Network Properties of Preterm and At-Term Neonates. Hum Brain Mapp. 2025;46.

20. Wang J, Dong Q, Niu H. The minimum resting-state fNIRS imaging duration for accurate and stable mapping of brain connectivity network in children. Sci Rep. 2017;7.

21. The WHO Multicentre Growth Reference Study (MGRS): Rationale, planning, and implementation. Food Nutr Bull. 2004;25:S3–S84.

22. Fishburn FA, Ludlum RS, Vaidya CJ, Medvedev A V. Temporal Derivative Distribution Repair (TDDR): A motion correction method for fNIRS. Neuroimage. 2019;184.

23. Iester C, Bonzano L, Biggio M, Cutini S, Bove M, Brigadoi S. Comparing different motion correction approaches for resting-state functional connectivity analysis with functional near-infrared spectroscopy data. Neurophotonics. 2024;11.

24. Gossé LK, Pinti P, Wiesemann F, Elwell CE, Jones EJH. Developing customized NIRS-EEG for infant sleep research: methodological considerations. Neurophotonics. 2023;10.

25. Guglielmini S, Chen Z, Wolf M. DL-QC-fNIRS: a deep learning tool for automated quality control in functional near-infrared spectroscopy signals. Neurophotonics [online serial]. SPIE; 2025;13:15001. Accessed at: 10.1117/1.NPh.13.1.015001.

26. Klein F. Optimizing spatial specificity and signal quality in fNIRS: an overview of potential challenges and possible options for improving the reliability of real-time applications. Frontiers in Neuroergonomics 2024.

27. Dans PW, Foglia SD, Nelson AJ. Data processing in functional near-infrared spectroscopy (Fnirs) motor control research. Brain Sci. 2021.

28. Kassab A, Toffa DH, Robert M, et al. Cortical Hemodynamics of Electrographic Status Epilepticus in the Critically Ill. Epilepsia. Epub 2024.

29. Wu C-W, Chen JJ, Lin CK, et al. Hemodynamics and Tissue Optical Properties in Bimodal Infarctions Induced by Middle Cerebral Artery Occlusion. Int J Mol Sci. Epub 2022.

30. Blanco B, Lloyd-Fox S, Begum-Ali J, et al. Cortical responses to social stimuli in infants at elevated likelihood of ASD and/or ADHD: A prospective cross-condition fNIRS study. Cortex. 2023;169.

31. Lanka P, Bortfeld H, Huppert TJ. Correction of global physiology in resting-state functional near-infrared spectroscopy. Neurophotonics. 2022;9.

32. Hong YM, Cho DH, Kim JK. Developmental outcomes of very low birth weight infants with catch-up head growth: a nationwide cohort study. BMC Pediatr. 2023;23.

33. Mayrink ML de S, Villela LD, Méio MDBB, et al. The trajectory of head circumference and neurodevelopment in very preterm newborns during the first two years of life: a cohort study. J Pediatr (Rio J). 2024;100.

34. Pedaveeti M, Iqbal F, Purkayastha J, Bharadwaj SK, Patil AK, Lewis LES. Comparative Growth Outcomes in Very Low Birth Weight Infants: Evaluating Different Feeding Strategies. Indian J Pediatr. 2025;92.

35. Miller TA, Zak V, Shrader P, et al. Growth Asymmetry, Head Circumference, and Neurodevelopmental Outcomes in Infants with Single Ventricles. Journal of Pediatrics. 2016;168.

36. Vinall J, Grunau RE, Brant R, et al. Slower postnatal growth is associated with delayed cerebral cortical maturation in preterm newborns. Sci Transl Med. 2013;5.

37. Neubauer V, Fuchs T, Griesmaier E, Kager K, Pupp-Peglow U, Kiechl-Kohlendorfer U. Poor postdischarge head growth is related to a 10% lower intelligence quotient in very preterm infants at the chronological age of five years. Acta Paediatrica, International Journal of Paediatrics. 2016;105.

38. Sammallahti S, Heinonen K, Andersson S, et al. Growth after late-preterm birth and adult cognitive, academic, and mental health outcomes. Pediatr Res. 2017;81.

39. Beunders VAA, Roelants JA, Suurland J, et al. Early Ultrasonic Monitoring of Brain Growth and Later Neurodevelopmental Outcome in Very Preterm Infants. American Journal of Neuroradiology. 2022;43.

40. Taine M, Forhan A, Morgan AS, et al. Early postnatal growth and subsequent neurodevelopment in children delivered at term: The ELFE cohort study. Paediatr Perinat Epidemiol. 2021;35.

41. Bartholomeusz HH, Courchesne E, Karns CM. Relationship Between Head Circumference and Brain Volume in Healthy Normal Toddlers, Children, and Adults. Neuropediatrics. Epub 2002.

42. Graaff JS, Roza SJ, Walstra AN, et al. Associations of Maternal Folic Acid Supplementation and Folate Concentrations During Pregnancy With Foetal and Child Head Growth: The Generation R Study. Eur J Nutr. Epub 2015.

43. Hindmarsh PC, Geary M, Rodeck CH, Kingdom J, Cole T. Factors Predicting Ante- And Postnatal Growth. Pediatr Res. Epub 2008.

44. Smyser CD, Inder TE, Shimony JS, et al. Longitudinal Analysis of Neural Network Development in Preterm Infants. Cerebral Cortex. Epub 2010.

45. Jakab A, Schwartz E, Kasprian G, et al. Fetal Functional Imaging Portrays Heterogeneous Development of Emerging Human Brain Networks. Front Hum Neurosci. Epub 2014.

46. Burton GJ, Fowden AL. The Placenta: A Multifaceted, Transient Organ. Philosophical Transactions of the Royal Society B Biological Sciences. Epub 2015.

47. Sharma D, Shastri S, Sharma P. Intrauterine Growth Restriction: Antenatal and Postnatal Aspects. Clin Med Insights Pediatr. Epub 2016.

48. Uberos J, Jiménez-Montilla S, Machado-Casas I, Láynez-Rubio C, Fernández-Marín E, Campos-Martínez A. The Association Between Restricted Intra-Uterine Growth and Inadequate Postnatal Nutrition in Very-Low-Birth-Weight Infants and Their Neurodevelopmental Outcomes: A 50-Month Follow-Up Study. British Journal of Nutrition. Epub 2021.

49. Arichi T, Fagiolo G, Varela M, et al. Development of BOLD Signal Hemodynamic Responses in the Human Brain. Neuroimage. Epub 2012.

50. Watanabe H, Shitara Y, Aoki Y, et al. Hemoglobin Phase of Oxygenation and Deoxygenation in Early Brain Development Measured Using fNIRS. Proceedings of the National Academy of Sciences. Epub 2017.

51. Cusack R, Wild CJ, Linke AC, Arichi T, Lee DSC, M. Han VK. Optimizing Stimulation and Analysis Protocols for Neonatal fMRI. PLoS One. Epub 2015.

52. Karen T, Morren G, Haensse D, Bauschatz A, Bucher HU, Wolf M. Hemodynamic Response to Visual Stimulation in Newborn Infants Using Functional Near-infrared Spectroscopy. Hum Brain Mapp. Epub 2007.

